# Copy Number Analysis in Congenital Nevi: Concordance and Diagnostic Limitations of aCGH, sWGS, and Methylation Profiling

**DOI:** 10.64898/2026.03.03.26347388

**Authors:** Anton Karelin, Ines B. Brecht, Michaela Pogoda, German Demidov, Michael Abele, Dominik T. Schneider, Daniel Aldea, Heather C Etchevers, Susana Puig, Matthias Hahn, Christopher Schroeder, Stephan Forchhammer, the MELCAYA consortium

## Abstract

**Background:** Distinguishing benign proliferative nodules (PNs) from melanoma arising within congenital melanocytic nevi remains a major diagnostic challenge. Copy number alteration (CNA) analysis is widely used to support classification, but current criteria were developed using array comparative genomic hybridization (aCGH). The performance of alternative platforms such as shallow whole-genome sequencing (sWGS) and methylation arrays in this setting is poorly defined.

**Objectives:** The objective of this study is to compare CNA profiles obtained from aCGH, sWGS, and methylation arrays in atypical nodules arising within congenital nevi, and to correlate these molecular findings with clinical outcomes.

**Methods:** Sixteen samples from fourteen patients were retrospectively analyzed using all three platforms. CNAs were cataloged, concordance across methods was quantified using the Jaccard index, and molecular classifications were compared. Clinical follow-up was reviewed to provide clinical context.

**Results:** aCGH detected 39 CNAs, sWGS 60, and methylation profiling 66. Concordance was highest between sWGS and methylation (mean Jaccard 0.67), followed by aCGH versus sWGS (0.64) and aCGH versus methylation (0.49). Cases with high aneuploidy demonstrated strong cross-platform agreement, whereas low-burden lesions exhibited greater variability between methods. Divergent molecular classifications were observed in six cases.

**Conclusions:** While all methods reliably detect broad chromosomal changes, sWGS and methylation arrays identify many additional focal CNAs that may not align with CGH-based diagnostic criteria. Until platform-specific thresholds are established, aCGH remains the most conservative and clinically validated approach for evaluating proliferative nodules in congenital nevi.

**Patient consent statement:** Written informed consent was given by all patients or their legal guardians. Consent was waived for deceased patients or samples older than 5 years.

**SIGNIFICANCE:** Accurate molecular classification of melanocytic proliferations in congenital nevi is essential but challenging, particularly in patients with multiple proliferative nodules. This study provides the first systematic comparison of aCGH, sWGS, and methylation-based CNA profiling in this setting. We show that higher-resolution platforms detect substantially more focal aberrations, which can lead to discordant and potentially overcalled malignancy assessments when applying CGH-derived criteria. Our findings highlight the need for platform-adapted diagnostic frameworks and support continued use of CGH as the most conservative and clinically validated method for risk stratification.

**GRAPHICAL ABSTRACT:** 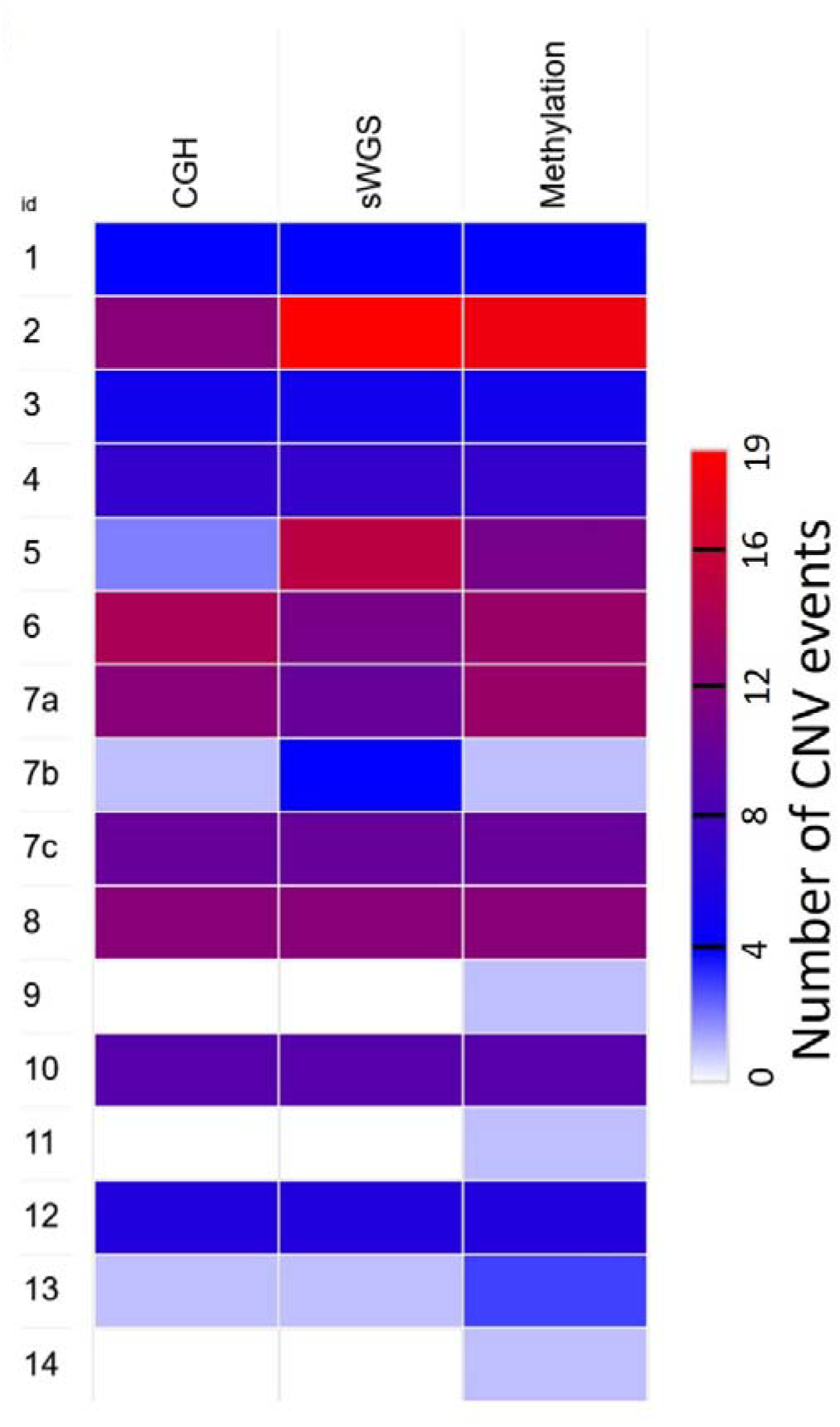

## 1. INTRODUCTION

Pediatric melanoma is a rare neoplasm, with an estimated incidence of four cases per million children and adolescents under 20 years of age annually ^1,2^. Incidence rises markedly during puberty and adolescence, where melanoma accounts for approximately 7% of all malignancies among individuals aged 15–19 years ^3^. Most cases occur in adolescents, whereas melanoma in children under 10 years remains exceptionally uncommon ^2,4^.

Pediatric melanomas comprise several biologically distinct subtypes, including Spitz melanomas, conventional melanomas, melanomas arising within large or giant congenital melanocytic nevi (CMN), and rare variants. Prognosis varies widely between these subgroups. Spitz melanocytomas and Spitz melanomas show a high rate of nodal involvement but low melanoma-specific mortality ^5-7^. In contrast, conventional melanomas and melanomas arising in congenital nevi behave more aggressively. Although rare, melanomas arising in CMN account for a disproportionate number of fatal pediatric melanoma cases, particularly in children under five years of age ^2,6,7^.

Diagnosis of melanoma arising in CMN remains challenging. Giant CMN may contain multiple, sometimes rapidly enlarging nodules, and distinguishing benign proliferative nodules (PN) from melanoma can be difficult based on clinical and histologic criteria alone. Histopathologic assessment typically evaluates mitotic activity, ulceration, cytologic atypia, and architectural features to differentiate PNs from melanoma ^2,8-10^.

Molecularly, PNs and melanomas differ mainly in their patterns of copy number alterations (CNAs). Benign PNs typically exhibit whole-chromosome gains or losses, whereas melanomas show multiple segmental CNAs, including partial chromosomal gains and deletions ^11-13^. Comparative genomic hybridization (CGH) is widely regarded as the reference standard for CNA analysis in this setting because it reliably discriminates between simple numerical aberrations and the complex segmental patterns typical of malignancy ^11-13^. However, array-CGH is technically demanding and limited to specialized laboratories.

Alternative CNA detection approaches have become increasingly available. Shallow whole-genome sequencing (sWGS) provides genome-wide CNA profiles at low sequencing depth and is suitable for FFPE tissue, with diagnostic sensitivity comparable to CGH in melanocytic tumors ^14,15^. CNAs can also be inferred from DNA methylation arrays using computational tools such as Conumee or Epicopy, enabling genome-wide CNA profiles from the same data used for methylation-based tumor classification ^16,17^.

Despite their growing use, no prior study has systematically compared sWGS- and methylation-derived CNA profiles with CGH specifically in proliferative nodules and melanomas arising within congenital nevi. Here, we compare CNA detection across all three platforms and correlate molecular findings with clinical outcome.

## 2. METHODS

### 2.1 Study cohort

Children and young adults (0–30 years) with atypical melanocytic proliferations arising within a congenital melanocytic nevus between 2005 and 2024 were included. Cases originated either from referral specimens evaluated for expert dermatopathological consultation or from patients treated at the University Hospital Tübingen. Archival FFPE samples were re-evaluated, and 14 of 28 eligible cases were included after excluding samples with insufficient material or DNA yield. Sixteen lesions from these 14 patients were analyzed, as one patient had three nodules excised at different time points. Written informed consent was obtained where applicable; archival cases older than five years were included under an institutional consent waiver.

### 2.2 Histology and molecular classification

Only lesions in which morphology alone did not allow reliable distinction between PN and melanoma were included. All cases were jointly assessed by at least two dermatopathologists specialized in melanocytic tumors. Molecular classification followed the criteria of Bastian et al. ^12^. Lesions without CNAs or with whole-chromosome gains/losses and a single non–melanoma-typical arm-level aberration were classified as benign. Single melanoma-typical segmental aberrations were considered borderline. Multiple (≥2) segmental or arm-level aberrations defined malignant molecular profiles.

### 2.3 Array comparative genomic hybridization

Comparative genomic hybridization (CGH) analyses were performed in two distinct settings. For cases 1–7a and 7b, CGH was newly conducted retrospectively. DNA was extracted from formalin-fixed, paraffin-embedded (FFPE) tissue samples, which were processed following standardized protocols for microdissection and DNA isolation. Microarray analyses were carried out using the SurePrint G3 Human CGH Microarray Kit 4×180K (Agilent Technologies, Santa Clara, USA). Hybridizations were scanned on an Agilent SureScan DX Microarray Scanner, with human reference DNA (female or male, Agilent Technologies) serving as the comparative reference. Data analysis was performed using Agilent Cytogenomics software (v5.1.2.1).

For cases 7c–14, existing CGH data from routine diagnostics, generated between 2009 and 2021, were utilized. Tumor tissue was manually microdissected from four to six 25-µm-thick paraffin sections, deparaffinized, and digested with proteinase K. DNA extraction and labeling were performed using genomic DNA universal linkage system labeling kits (Agilent Technologies, Santa Clara, USA). Hybridization employed the Agilent Human Genome CGH 105K microarray (Agilent Technologies). Arrays were scanned using an Agilent DNA microarray scanner, and image data were analyzed with Feature Extraction software (v10.5.1.1), DNA Analytics (v4.0.85), and Genomic Workbench Lite Edition (v6.5.0.18).

### 2.4 sWGS

DNA was extracted from formalin-fixed, paraffin-embedded (FFPE) tissue samples, which were processed following standardized protocols for microdissection and DNA isolation. The DNA was quantified using the Qubit Fluorometer and dsDNA High sensitivity assay (both Thermo Fisher Scientific, Waltham, USA). The genomic fragment lengths of the samples were assessed with the Tapestation 4200 and the Genomic DNA ScreenTape Analysis (Agilent Technologies, Santa Clara, USA). Since some of the DNA samples showed discolorations, all of them were cleaned up with paramagnetic beads (SPRISelect, Beckman Coulter, Brea, USA) to reduce potential contaminants. 100 ng of the DNA were subsequently sheared and prepared for Next Generation Sequencing (NGS) using the NEBNext UltraShear FFPE DNA Library Prep Kit with NEBNext Unique Dual Index UMI Adaptors (both New England Biolabs, Ipswich, USA) with slight adjustments to the fragmentation incubation and the PCR protocol to accommodate for differences in the original sample qualities. The concentrations of the libraries were assessed using the Qubit Fluorometer and dsDNA High Sensitivity Assay-Kit (both Thermo Fisher Scientific, Waltham, USA), while fragment sizes were determined with the Fragment Analyzer 5300 and the Fragment Analyzer DNA HS NGS fragment Fragment kit (Agilent Technologies, Santa Clara, USA).

Sequencing was performed on a NovaSeq 6000 (Illumina, San Diego, CA, USA) in paired-end mode (2×154 bp) with an average of 177 million reads per sample.

### 2.5 Methylation

DNA was extracted from formalin-fixed, paraffin-embedded (FFPE) tissue samples, which were processed following standardized protocols for microdissection and DNA isolation. The DNA was quantified using the Qubit Fluorometer and dsDNA High sensitivity assay (both Thermo Fisher Scientific, Waltham, USA). The genomic fragment lengths of the samples were assessed with the Tapestation 4200 and the Genomic DNA ScreenTape (both Agilent Technologies, Santa Clara, USA).

200 ng of DNA were prepared using the Twist Targeted Methylation Sequencing Workflow (Twist Bioscience, South San Francisco, USA) including the NEBNext Enzymatic Methyl-seq Library Preparation Protocol (New England Biolabs, Ipswich, USA) according to the manufacturers’ protocol with 8-plex pre-capture pooling and targeted enrichment using the Twist Human Methylome enrichment panel. The concentrations of the libraries and pools were assessed using the Qubit Fluorometer and dsDNA High Sensitivity Assay-Kit (both Thermo Fisher Scientific, Waltham, USA), while fragment sizes were determined with the TapeStation 4200 and the High Sensitivity D1000 ScreenTape (both Agilent Technologies, Santa Clara, USA).

Sequencing was performed on a NovaSeq 6000 (Illumina, San Diego, CA, USA) in paired-end mode (2×154 bp) with an average of 227 million reads per sample.

### 2.6. Sequencing

Shallow whole-genome sequencing (sWGS) and methylation sequencing (EM-seq) data as well as quality control were analyzed with an in-house pipeline called megSAP (version archived at Zenodo: https://zenodo.org/records/15063428). Sequencing reads were mapped to human genome reference GRCh38 using BWA-MEM2 for sWGS and Bismark for EM-seq ^18,19^.

Mean coverage was 6.78x (range: 5.57-7.62) and 78.01x (range: 19.48-137.31) for sWGS and EM-seq respectively.

### 2.7. Copy-Number Analysis

Copy-number alteration (CNA) analysis from sWGS and EM-seq data was performed using ClinCNV, a segmentation-based algorithm optimized for low-coverage whole-genome and targeted sequencing data ^20^. As no matched normal samples were available, CNA calling was performed using a tumor-only approach, assuming diploidy across the majority of the genome.

Coverage tracks were generated at 10 kb resolution, and GC correction was applied. CNA detection was executed with tumor-only mode using the following parameters:

--onlyTumor, --scoreS 1000, --lengthS 10, --minimumNumOfElemsInCluster 15, --purityStep 5, -- clonePenalty 1000, --minimumPurity 5, --filterStep 0, with hg38 as the reference genome. Pseudoautosomal regions on chromosome X were excluded from the analysis to avoid sex-related copy number bias.

All detected CNA profiles were assessed by manual inspection. CNAs were categorized as whole chromosome or chromosome arm gains and deletions.

### 2.8. Statistics

Statistical analyses were performed using Microsoft Excel (version 16.0). Descriptive statistics were calculated as appropriate. Concordance between CNA profiles obtained by aCGH, sWGS, and methylation arrays was quantified using the Jaccard similarity coefficient, defined as the ratio of shared to total aberrations for each method pair. Heatmap visualizations were generated using Morpheus (Broad Institute; https://software.broadinstitute.org/morpheus/).

### 2.9. Ethics statement

The study was approved by the Ethics Committee of the University of Tübingen (Project ID 399/2023BO1) and conducted in accordance with the Declaration of Helsinki.

## 3. RESULTS

### 3.1. Epidemiology, clinical and histological presentation

The cohort included 14 patients aged 8 days to 26 years (nine males, five females). Most lesions arose within giant CMN located on the back or trunk. Two cases involved congenital blue nevi and two involved medium- or small-sized CMN with atypical features. Histopathologically, most nodules represented proliferative nodules (PN), including spitzoid variants requiring molecular workup for classification. Follow-up ranged from 2 to 94 months. Three patients died from their disease, seven remained disease-free, and four lacked follow-up (Table 1).

**Table 1.** Clinical and histopathological characteristics of the study cohort. **Abbreviations:** CMN, (giant unless otherwise specified) congenital melanocytic nevus; PN, proliferative nodule; d, days; mo, months; y, years; D, deceased; A, alive without disease; n.a., not available.

| Case | Age | Sex | Clinical | Histology | Follow-up |
| --- | --- | --- | --- | --- | --- |
| 1 | 12- 17 y | m | Nodule in CMN, head/neck | PN on CMN | 82 mo, D |
| 2 | 0-5 y | m | CMN, scalp | PN on CMN | 14 mo, D |
| 3 | 0-5 y | f | Nodule in CMN, right hip | PN on CMN | 17 mo, A |
| 4 | 0-5 y | f | Dark pigmented nodule in CMN, right shoulder | PN on CMN | 48 mo, A |
| 5 | 12- 17 y | m | Medium-sized CN with nodular areas on the back of the left hand | Atypical spitzoid<br>PN on CMN | 87 mo, A |
| 6 | 0-5 y | f | CMN with multiple nodular areas, buttock | PN on CMN | 49 mo, A |
| 7a | 0-5 y | f | CMN with multiple nodular areas, back | PN on CMN | 2 mo, D (after excision of the lymph node metastasis) |
| 7b | 0-5 y |  | CMN with multiple nodular areas, back | PN on CMN |  |
| 7c | 0-5 y |  | Nodular swelling, left axilla | lymph node metastasis |  |
| 8 | 0-5 y | f | CMN with multiple nodular areas, back | PN on CMN | 36 mo, A |
| 9 | 0-5 y | f | CMN with darker areas, right shoulder | PN on CMN | 26 mo, A |
| 10 | 0-5 y | m | CMN with nodular areas, back | PN on CMN | n.a. |
| 11 | 6-11 y | f | n.a. | PN on congenital blue nevus | n.a. |
| 12 | 12- 17 y | m | n.a. | PN on CMN | 94 mo, A |
| 13 | 6-11 y | m | n.a. | PN on large congenital blue nevus | n.a. |
| 14 | 24-29 y | m | Small, papillomatous CMN | Spitzoid PN on CMN | n.a. |

### 3.2 Comparative Analysis of Copy Number Alteration Detection by aCGH, sWGS, and Methylation Profiling

All lesions were analyzed by aCGH, sWGS, and methylation-derived CNA profiling, and all detected aberrations are listed in Table 2. Across the cohort, the platforms revealed a broadly consistent pattern of chromosomal gains and losses, but with marked differences in the number and resolution of detected events. aCGH identified 39 CNAs, whereas sWGS and methylation profiling detected 60 and 66, respectively. Despite this, substantial overlap existed: 34 CNAs were shared between aCGH and sWGS, 34 between aCGH and methylation, and 45 between sWGS and methylation. A core set of 32 CNAs was concordantly detected across all three methods.

**Table 2.** Copy number alterations detected by aCGH, sWGS, and methylation-based CNA profiling across all cases. For each lesion, all chromosomal gains and losses identified by array comparative genomic hybridization (aCGH), shallow whole-genome sequencing (sWGS), and methylation-derived CNA analysis are listed. **Abbreviations:** del, deletion; gain, copy number gain; qter, terminal end of the long arm; pter, terminal end of the short arm; none, no detectable CNAs.

| Case | CGH | sWGS | Methylation |
| --- | --- | --- | --- |
| 1 | del(5, 7, 10, 12) | del(5, 7, 10, 12) | del(5p, 7, 10q, 12q) |
| 2 | gain(1q, 13q12.2–q32.1); del(3, 5p10–p12, 5q12.1–q14.1, 9, 10, 11, 12, 14, 17, 21) | gain(1, 2, 4, 6p, 7q, 13q12.12–q21.32, 13q22.1–q32.1, 15, 22); del(3, 5p10–p12, 5q12.1–q35.1, 9, 10, 11, 12, 14, 17, 21) | gain(1, 2, 4, 6p, 7, 13q12.12–q21.32, 13q22.1–q32.1, 15, 22); del(3p, 5q12.1–q14.1, 9, 10, 11p, 12, 14, 17, 21) |
| 3 | gain(2, 8, 13, 20, 22) | gain(2, 8, 13, 20, 22) | gain(2, 8, 13, 20, 22) |
| 4 | gain(6, 8, 9, 16, 18, 19, 20) | gain(6, 8, 9, 16, 18, 19, 20) | gain(6, 8, 9, 16, 18, 19, 20) |
| 5 | del(4q22.2–q28.2, 9) | gain(1p22.2–p36.13, 3p12.3–p25.3, 3q11.2–qter, 5p12–p13.3, 5q, 8q12.1–q24.23, 11q, 12q12–q23.3, 15q12–q25.2); del(1p36.13–pter, 2, 4q22.2–q28.2, 6q12–q22.31, 9, 16p) | gain(3p12.3–p25.3, 3q11.2–qter, 5p12–p15.3, 5q, 11p11.2–p15.4, 11q, 12q12–q23.3); del(1p36.13–pter, 4q22.2–q28.2, 9, 16) |
| 6 | gain(4, 8, 9, 12, 13, 14, 16, 19, 20, 21, 22); del(5, 10, 18) | gain(4, 8, 9, 12, 13, 14, 16q11.2–q24.1, 21); del(5, 10, 18) | gain(4, 9, 12, 13, 14, 16q11.2–q24.1, 19q13.31–q13.33, 21); del(1p32.2–pter, 5, 6p12.2–p22.2, 10, 18) |
| 7a | gain(7, 8, 9, 12, 13, 16q, 17, 19, 20, 22); del(5, 10) | gain(7, 8, 9, 12, 13, 16q, 17q, 22); del(5, 10) | gain(7, 8, 9, 12, 13, 16p11.2–pter, 16q12.2–qter, 17, 19p13.11–pter, 19q13.11–q13.33, 22); del(5, 10) |
| 7b | del(7) | gain(5q11.1–q33.1); del(7, 8q11.1–q23.3, 16p12.1–p13.3) | del(7) |
| 7c | gain(1q, 7, 8, 20); del(3q13.3–qter, 5, 11, 16q, 17, 18) | gain(1q, 7, 8, 20); del(3q12.2–qter, 5, 11, 16q, 17, 18) | gain(1q, 7, 8, 20); del(3q12.2–qter, 5, 11, 16q, 17, 18) |
| 8 | gain(1q, 8, 16, 20); del(3, 5, 7, 11, 12, 14, 19, 21) | gain(1q, 8, 16, 20); del(3, 5, 7, 11, 12, 14, 19, 21) | gain(1q, 8, 16, 20); del(3, 5, 7, 11, 12, 14, 19, 21) |
| 9 | none | none | del(20q11.21–qter) |
| 10 | gain(7, 8, 12, 13, 16, 18, 20, 21); del(3) | gain(7, 8, 12, 13, 16, 18, 20, 21); del(3) | gain(7, 8, 12, 13, 16, 18, 20, 21); del(3) |
| 11 | none | none | del(8q) |
| 12 | gain(5, 15, 18, 20, 21, 22) | gain(5, 15, 18, 20, 21, 22) | gain(5, 15, 18, 20, 21, 22) |
| 13 | gain(6p21.1–pter) | gain(6p22.2–pter) | gain(6p22.2–pter); del(8q11.1–q24.23, 13q12.13–q33.3) |
| 14 | none | none | del(20q11.21–qter) |

As expected, aCGH predominantly captured broad whole-chromosome alterations, while sWGS and methylation profiling revealed numerous additional subchromosomal segmental and focal events. Methylation profiling, in particular, identified recurrent terminal and arm-level deletions (e.g., del(20q11.21–qter), del(8q)) that were not detected by either aCGH or sWGS, suggesting increased sensitivity to focal or possibly subclonal aberrations. Jaccard similarity coefficients demonstrated the highest concordance between sWGS and methylation profiling (mean 0.67), followed by aCGH versus sWGS (0.64) and aCGH versus methylation (0.49). Cases with extensive aneuploidy—such as cases 3, 4, 7c, 8, 10, and 12—showed perfect concordance across all methods, whereas cases with few aberrations exhibited greater variability, largely driven by additional focal events detected by methylation arrays.

To visualize CNA burden across the cohort, a heatmap summarizing the number of CNAs per case and platform was generated, together with representative CNA profiles illustrating platform-specific resolution differences (Figure 1). Beyond the raw CNA counts, we assessed how platform-specific variation influenced molecular classification (Table 3). Based on established CGH-derived criteria, where lesions with no CNAs or exclusively whole-chromosome gains or losses are considered benign, and profiles with two or more segmental or arm-level aberrations are classified as malignant, aCGH classified most lesions as benign (13/16), whereas sWGS and methylation profiling resulted in malignant classifications in 5/16 and 7/16 cases, respectively. Six cases showed complete concordance across all platforms, four showed minor discrepancies, and six displayed major discrepancies that shifted the diagnostic category. Minor discrepancies were defined as differences in the number or type of detected CNAs that did not alter the resulting molecular classification, whereas major discrepancies were defined as discordances that shifted the diagnostic category (benign, borderline, or malignant). These divergences were primarily driven by additional segmental or focal CNAs detected by sWGS or methylation profiling, underscoring the challenge of applying CGH-based criteria to higher-resolution platforms.

**Table 3.**
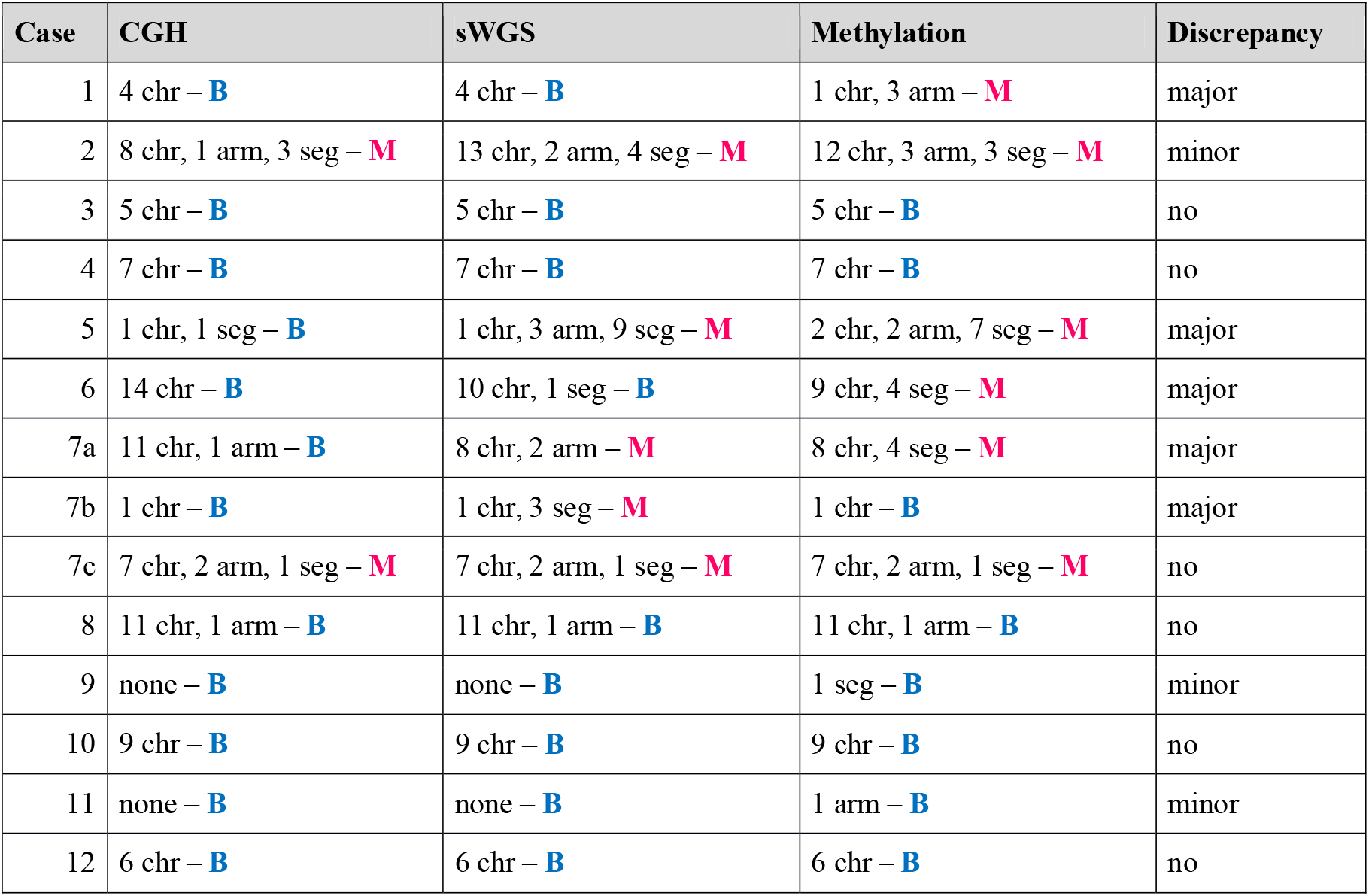
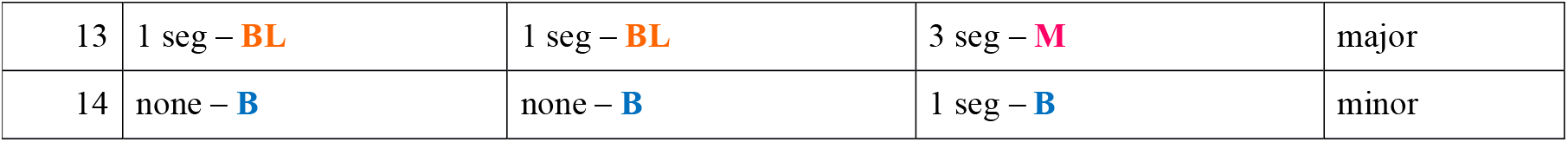
Molecular classifications based on CNA patterns detected by aCGH, sWGS, and methylation profiling. Shown are the numbers of whole-chromosome (chr), chromosome-arm (arm), and segmental (seg) alterations per platform, the resulting molecular classification (B = benign, BL = borderline, M = malignant), and the degree of discrepancy between methods. **Abbreviations:** chr – chromosome; arm – chromosome arm; seg – segment; aCGH – array comparative genomic hybridization; sWGS – shallow whole-genome sequencing; B – benign; BL – borderline; M – malignant.

**Figure 1:**
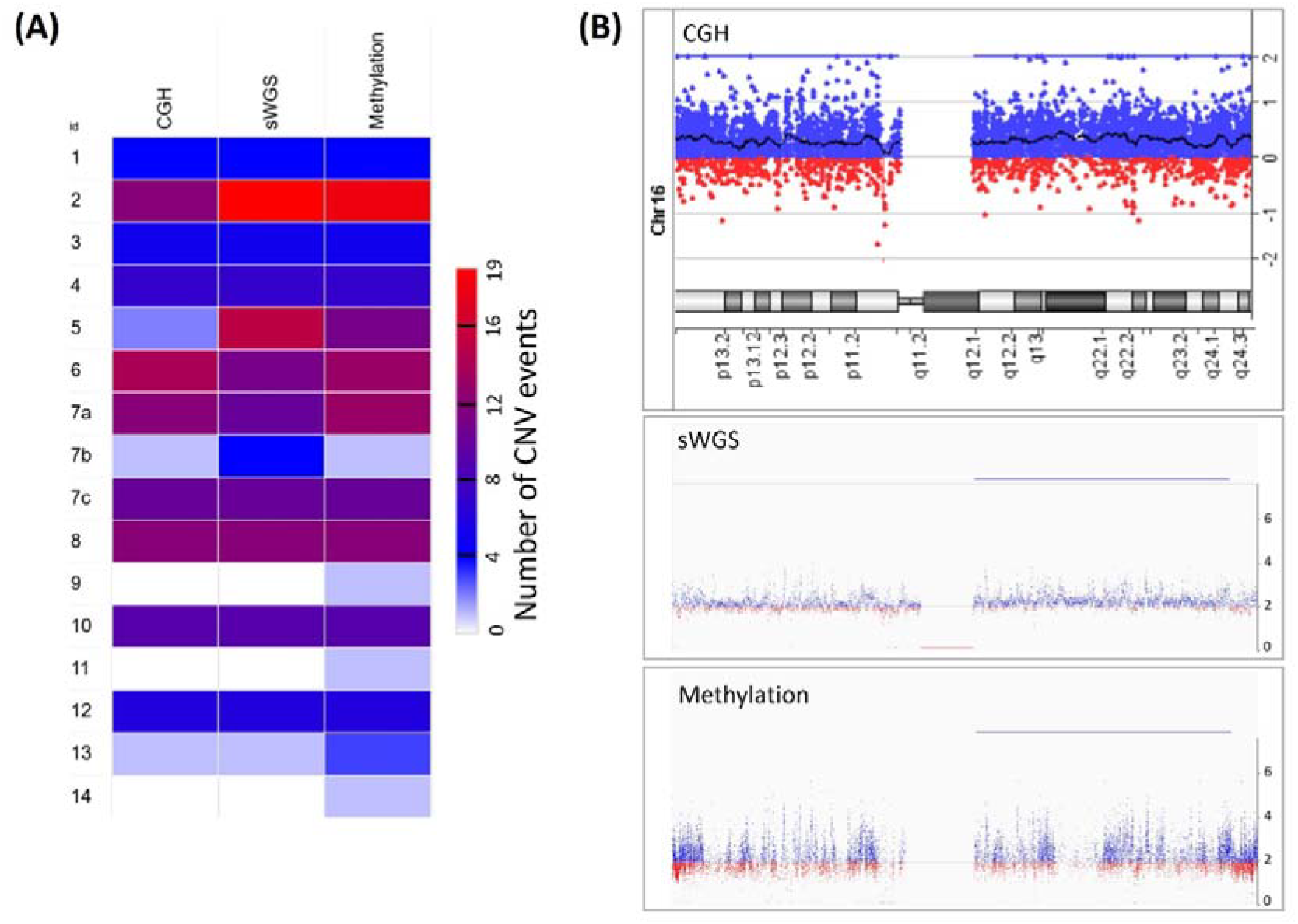
Cross-platform comparison of CNV detection by aCGH, sWGS, and methylation profiling. (**A**) Heatmap showing the number of chromosomal, arm-level, and segmental CNVs detected per case across all three platforms. High-burden cases demonstrate strong cross-platform concordance, whereas low-burden lesions display additional focal alterations, predominantly detected by methylation arrays. (**B**) Exemplary CNA profiles of chromosome 16 from case 6. While aCGH identifies a whole-chromosome gain, both sWGS and methylation profiling detect only a long segmental gain on 16q and do not call the gain on 16p, illustrating platform-specific differences in resolution and segmentation. CGH data are displayed on a log_2_ ratio scale, whereas for sWGS and methylation profiling the y-axis represents estimated absolute copy number on a linear scale, with 2 corresponding to diploidy.

### 3.3 Correlation of clinical outcome

Clinical follow-up was reviewed to assess the relationship between molecular classifications and patient outcomes. Interpretation was limited in patients with giant congenital nevi containing multiple proliferative nodules, as the excised lesion might not represent the biologically relevant precursor of metastasis.

This issue was most evident in patient 7, from whom three distinct nodules were sampled over time (cases 7a, 7b, and 7c). Cases 7a and 7b showed differing CNA patterns across platforms but were classified as benign by aCGH (and by methylation for 7b). In contrast, case 7c represented the lymph node metastasis and exhibited a highly complex CNA profile characteristic of melanoma. Neither 7a nor 7b shared the CNA pattern of the metastatic lesion, suggesting that the true primary melanoma node was not sampled. Thus, the molecular classifications of 7a and 7b cannot be directly correlated with the patient’s adverse clinical course; these findings suggest they represent independent events.

Other discordant cases showed similar diagnostic uncertainty. In case 1, aCGH and sWGS classified the lesion as benign, whereas methylation profiling detected arm-level deletions consistent with a malignant profile. The patient ultimately died; however, due to the multifocal nature of the congenital nevus, it remains unclear whether the analyzed nodule represented the lesion that gave rise to metastasis. In contrast, cases 5 and 6 showed benign CNA profiles by aCGH but malignant-appearing profiles by sWGS and methylation analysis; both patients remained disease-free. Case 13 showed borderline results by aCGH and sWGS but a malignant CNA profile derived from methylation sequencing; no clinical follow-up was available.

Across the cohort, three patients experienced fatal outcomes. Because several of these patients had multiple nodules and the true primary melanoma focus often could not be unequivocally identified, formal performance metrics such as sensitivity and specificity could not be reliably established at the lesion level. Qualitatively, aCGH tended to classify fewer lesions as malignant than sWGS and methylation profiling, particularly in patients with an overall benign clinical course, whereas the higher-resolution platforms more often yielded malignant CNA patterns, including in lesions from patients who remained disease-free. These observations highlight both the increased sensitivity of sWGS and methylation-based CNA analysis and the difficulty of attributing biological significance to small focal aberrations in multifocal congenital nevi where the causative melanoma precursor may remain unsampled.

## 4. DISCUSSION

Although aCGH, sWGS, and methylation-based CNA analysis have often been described as comparable approaches for the detection of larger CNAs, our study demonstrates that relevant differences emerge when these methods are applied to atypical melanocytic proliferations in congenital nevi. Previous studies have reported high concordance between aCGH and next-generation sequencing (NGS)–based approaches for detecting broad CNAs in well-characterized genomic regions ^21-25^. In contrast, we observed substantial variability in the detection of segmental and focal aberrations, which in several cases resulted in divergent molecular classifications. These discrepancies likely reflect differences in technical resolution, genomic coverage, and analytical sensitivity across the platforms ^21-25^.

Interpreting molecular findings in this clinical setting is inherently challenging, particularly in patients with giant congenital nevi who frequently harbor multiple proliferative nodules. In such cases, it is often unclear whether the sampled lesion corresponds to the biologically relevant precursor of metastasis. This issue has been repeatedly emphasized in the literature ^9,11^ and was illustrated most clearly in our cohort by patient 7. This aligns with prior reports noting that melanomas arising in congenital nevi often develop within complex, heterogeneous nodular fields where histology and molecular profiles may diverge ^2,9,11,12^.

In this context, it is important to emphasize that current diagnostic criteria for molecular classification in congenital nevi and proliferative nodules were developed using CGH ^9,12-14^. Numerical aberrations involving whole chromosomes are considered typical for benign proliferative nodules, while melanomas are characterized by multiple segmental aberrations ^2,9,11,12^. Because these criteria were validated in cohorts assessed by CGH, they may not directly translate to higher-resolution platforms. sWGS and methylation profiling exhibit increased sensitivity for detecting small CNAs or subclonal aberrations (<100 kb), particularly in FFPE tissue ^16,26-28^. While these additional findings might capture biologically meaningful alterations, they might also represent low-level or focal changes without prognostic significance, raising the possibility of diagnostic overinterpretation ^16,26-28^. In the other direction, in tissue sections selected for DNA extraction from ambiguous neoplasms, anomalous mosaic clones may be present at levels too low to meet detection thresholds, a limitation that has been described for both array-based and low-pass sequencing approaches^29,30^.

Our findings therefore support the need for platform-adapted diagnostic frameworks. High-resolution platforms such as sWGS and methylation arrays provide technical advantages—including improved genomic resolution and the ability to extract CNA data from the same sample used for methylation classification—but their increased sensitivity must be balanced against the risk of upstaging otherwise benign lesions. Larger, prospectively collected cohorts including unequivocally benign proliferative nodules and melanomas arising within congenital nevi will be essential to delineate which CNA patterns detected by higher-resolution methods carry true biological and prognostic relevance. Targeted validation with orthogonal techniques like digital PCR, FISH or MLPA, or deeper coverage with targeted NGS will be necessary to establish thresholds of reliability.

This study has several important limitations. First, the cohort size is inherently small due to the rarity of proliferative nodules and melanomas arising in congenital nevi, which restricts the statistical power of the analysis. Second, many patients had multiple nodules, and in some cases—including patient 7—the exact melanoma precursor could not be identified, preventing definitive lesion-level outcome correlation. Third, inherent differences between the platforms—including resolution, noise characteristics, and algorithmic approaches—likely contributed to discordances. Finally, the retrospective use of archival FFPE samples introduces variability in DNA quality and tumor content, which may influence CNA detection.

In summary, while all three platforms reliably captured broad chromosomal alterations, sWGS and methylation arrays identified substantially more focal and segmental CNAs. These differences significantly influenced molecular classification and must be interpreted cautiously, particularly in the setting of multinodular congenital nevi where the melanoma precursor may remain unsampled. Until platform-specific diagnostic criteria are established and supported by larger outcome-linked cohorts, CGH remains the most conservative and clinically validated approach for risk stratification in proliferative nodules arising within congenital nevi.

## Data Availability

The data presented in this study are available on request from the corresponding author.

## Acknowledgments

The findings and conclusions in this report are those of the authors and do not necessarily represent the official position of the European Commission.

The study was funded by an EU Horizon Grant (Melanoma in Childhood, Adolescents, and Young Adults -MELCAYA, HORIZON-MISS-2021-CANCER-02, ref. 101096667).

## Notes

NGS methods were performed with the support of the DFG-funded NGS Competence Center Tübingen (INST 37/1049–1), funded by the Deutsche Forschungsgemeinschaft (DFG, German Research Foundation) – Project-ID 286/2020B01—428994620. The STEP registry is funded by the Deutsche Kinderkrebsstiftung.

**Conflict of interest disclosure:** AK: None IBB: None MP: None GD: None MA: None DTS: None DA: None HCE: None SP: None MH: None CS: CS reports institutional grants from Illumina Inc. and Oxford Nanopore Technologies and research grants from BMS Stiftung Immunonkologie and Westdeutsche Studiengruppe GmbH outside the submitted work. SF: SF received personal honoraria from Kyowa Kirin, Stemline and Recordati Rare Diseases (speaker’s honoraria, advisory board), as well as institutional grants from BioNTech SE, Neracare and SkylineDX. All outside the submitted work.

### Competing Interest Statement

AK: None
IBB: None
MP: None
GD: None
MA: None
DTS: None
DA: None
HCE: None
SP: None
MH: None
CS: CS reports institutional grants from Illumina Inc. and Oxford Nanopore Technologies and research grants from BMS Stiftung Immunonkologie and Westdeutsche Studiengruppe GmbH outside the submitted work.
SF: SF received personal honoraria from Kyowa Kirin, Stemline and Recordati Rare Diseases (speaker's honoraria, advisory board), as well as institutional grants from BioNTech SE, Neracare and SkylineDX. All outside the submitted work.

### Funding Statement

This publication is funded by the European Union through the Horizon Research and Innovation program under Grant Agreement No 101096667.
NGS methods were performed with the support of the DFG-funded NGS Competence Center Tuebingen (INST 37/1049 1), funded by the Deutsche Forschungsgemeinschaft (DFG, German Research Foundation) Project ID 286/2020B01 428994620.
The STEP registry is funded by the Deutsche Kinderkrebsstiftung.

### Author Declarations

The study was approved by the Ethics Committee of the Medical Faculty of the University of Tuebingen (Project ID 399/2023BO1) and conducted in accordance with the Declaration of Helsinki.

### Summary of Updates

Corrected citation errors in references.

